# Real-World Data for Predicting Rapid Relapse Triple Negative Cancer: A Study Using NCDB and EHR Data

**DOI:** 10.64898/2026.01.28.26345096

**Authors:** Pallavi Jonnalagadda, Samilia Obeng-Gyasi, Daniel G. Stover, Barbara L. Andersen, Saurabh Rahurkar

## Abstract

**Background:** Many patients with triple-negative breast cancer (TNBC), particularly those who are older, Black, or insured by Medicaid, do not receive guideline-concordant treatment, despite its association with up to 4x higher survival. Early identification of patients at risk for rapid relapse may enable timely interventions and improve outcomes. This study applies machine learning (ML) to real-world data to predict risk of rapid relapse in TNBC.

**Methods:** We trained various ML models (logistic regression, decision trees, random forests, XGBoost, naïve Bayes, support vector machines) using National Cancer Database (NCDB) data and fine-tuned them using electronic health record (EHR) data from a cancer registry. Class imbalance was addressed using synthetic minority oversampling technique (SMOTE). Model performance was evaluated using sensitivity, specificity, positive predictive value (PPV), negative predictive value (NPV), receiver operating characteristics area under the curve ROC AUC, accuracy, and F1 scores. Transfer learning, cross-validation, and threshold optimization were applied to enhance the ensemble model’s performance on clinical data.

**Results:** Initial models trained on NCDB data exhibited high NPV but low sensitivity and PPV. SMOTE and hyperparameter tuning produced modest improvements. External testing on EHR data from a cancer registry had similar model performance. After applying transfer learning, cross-validation, and threshold optimization using the clinical data, the ensemble model achieved higher performance. The optimized ensemble model achieved a sensitivity of 0.87, specificity of 0.99, PPV of 0.90, NPV of 0.98, ROC AUC of 0.99, accuracy of 0.98, and F1-score of 0.88. This optimized model, leveraging readily available clinical data, demonstrated superior performance compared to initial NCDB-trained models and those reported in extant literature.

**Conclusions:** Transfer learning and threshold optimization effectively adapted ML models trained on NCDB data to an independent real-world clinical dataset from a single site, producing a high-performing model for predicting rapid relapse in TNBC. This model, potentially translatable to fast health interoperability resources (FHIR)-compatible workflows, represents a promising tool for identifying patients at high risk. Future work should include prospective external validation, evaluation of integration into clinical workflows, and implementation studies to determine whether the model improves care processes such as timely patient navigation and treatment planning.

**Author Summary:** In this study, we set out to understand which patients with triple-negative breast cancer might experience a rapid return of their disease. Many people with this aggressive form of cancer do not receive the treatments that are known to improve survival, especially patients who are older, Black, or insured through public programs. Being able to identify those at highest risk early in their care could help health teams provide timely support and ensure that patients receive the treatments they need.

To do this, we used information from a large national cancer database to build computer-based models that learn from patterns in patient data. We then refined these models using real medical records from a cancer center to make sure they worked well in everyday clinical settings. After adjusting and improving the models, we developed a tool that can correctly identify most patients who are likely to have a rapid return of their cancer.

Our hope is that this type of tool could eventually be built into routine care and help guide timely follow-up, support services, and treatment planning. More testing in real clinical environments will be important to understand how well the tool improves care and outcomes for patients.

## Introduction

Triple negative breast cancer (TNBC)—defined by the absence of estrogen (ER), progesterone (PR), and human epidermal growth factor-2 (HER2) receptors(1)—accounts for 10-15% of all breast cancers but accounts for approximately 40% of breast cancer-related deaths.(2–4) Compared to other subtypes, TNBC is more aggressive, less responsive to targeted therapies, and more likely to recur. Among patients with TNBC, about 15% of patients experience an especially aggressive course, known as rapid relapse TNBC (rrTNBC), where recurrence or cancer-related death occurs within 24 months of diagnosis.(5) These patients are resistant to standard therapies and demonstrate rapid progression of disease. Emerging evidence from the National Cancer Database (NCDB) suggests that the risk of rapid relapse is higher—and disproportionately affects—patients who are older, have public insurance, lower income, or live in rural areas.(6) These same groups are also less likely to receive guideline-concordant treatment, which typically includes both surgery and chemotherapy.

Importantly when guideline-concordant treatment is received by these patients, there is a 3x to 4x lower risk of rapid relapse.(6) These differences may reflect underlying challenges such as delayed or forgone care due to financial hardship, poor functional status, or misperceptions about the risks and benefits of treatment. Thus, early identification of patients at risk of rrTNBC may be critical in improving outcomes related to relapse and mortality among these patients.

Precision cancer medicine uses individualized information to inform better clinical decisions and often employs machine learning (ML) methods.(7) While ML approaches have been used to distinguish TNBC and non-TNBC,(8–13) few studies have examined

ML in the context of predicting relapse in TNBC.(14–16) In a study of 1,570 breast cancer patients, an ML algorithm predicted five-year mortality after discharge with a maximum accuracy of 77%, using laboratory measures such as platelet count, platelet-to lymphocyte ratio, age, lymphocyte-monocyte-ratio, and white blood cell count.(14) Another study leveraging multi-omics data for predicting rapid relapse demonstrated only modest predictive ability.(5) Similarly, a model distinguishing rapid relapse versus late relapse TNBC, achieved an AUC of 0.77. Thus, ML techniques have demonstrated significant promise in guiding treatment planning by identifying risk of relapse in patients with TNBC. Indeed, early identification of high-risk patients, who often belong to vulnerable populations, (6, 16, 17) may have potential benefits such as improved delivery of timely guideline-concordant treatment, and reduction in mortality, although extant studies focus on risk prediction rather than timing of relapse.

Provided at the point-of-care, such a screening tool leveraging socio-demographic, community needs factors, and clinical data from the EHR could enable early identification of patients at risk for rapid relapse, facilitating timely interventions to prevent disease progression and improve access to multidisciplinary care. Therefore, the objective of this study is to develop the rapid relapse predictive model (rrPM) by leveraging data from the NCDB and next testing it on real-world data (RWD) from the Ohio State University (OSU) Cancer registry. Findings from this study may inform the development of point of care clinical decision support (CDS) tools for oncologists.

### Ethics Approval

The Ohio State University Office of Responsible Research Practices deemed this study institutional review board exempt.

## Results

Of the 171,942 patients with TNBC in the NCDB, a total of 47,048 patients with TNBC from the NCDB met inclusion criteria and did not have missing data. Most patients with TNBC (Table 1) were between 45 – 64 years old (56.2%), identified as White (71.5%), and had private insurance or managed care (51.8%). Almost half the patients lived in neighborhoods where more than 10.9% of the population had less than high-school degrees (49.5%) and median incomes were <$50,353 (41.9%). Most patients with TNBC had low CCI scores of 0 (80.6%), had stage I-II disease (81.5%), and poorly differentiated tumors (85.1%). Overall, 14.5% had rapid relapse.

**Table 1:**
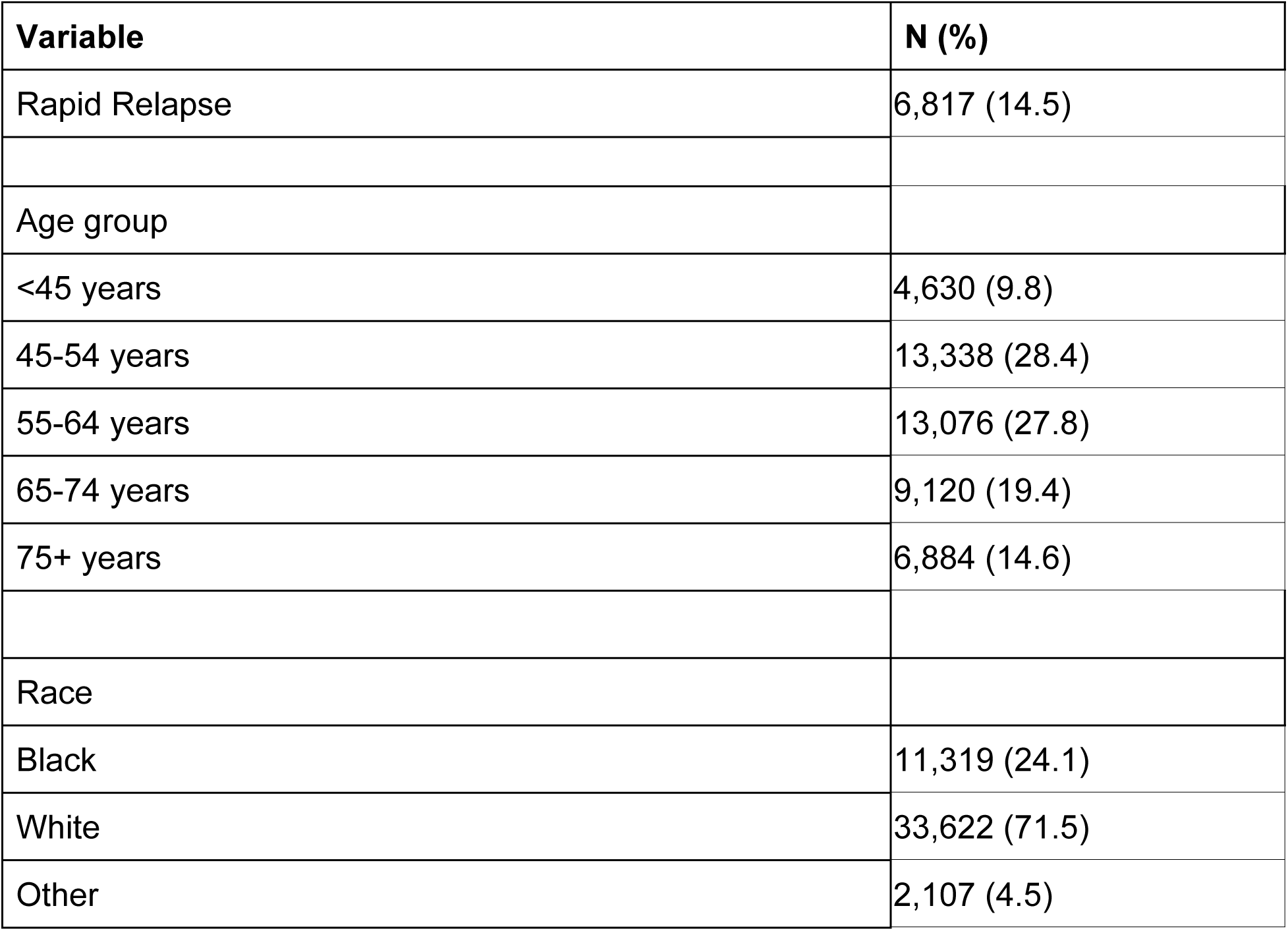
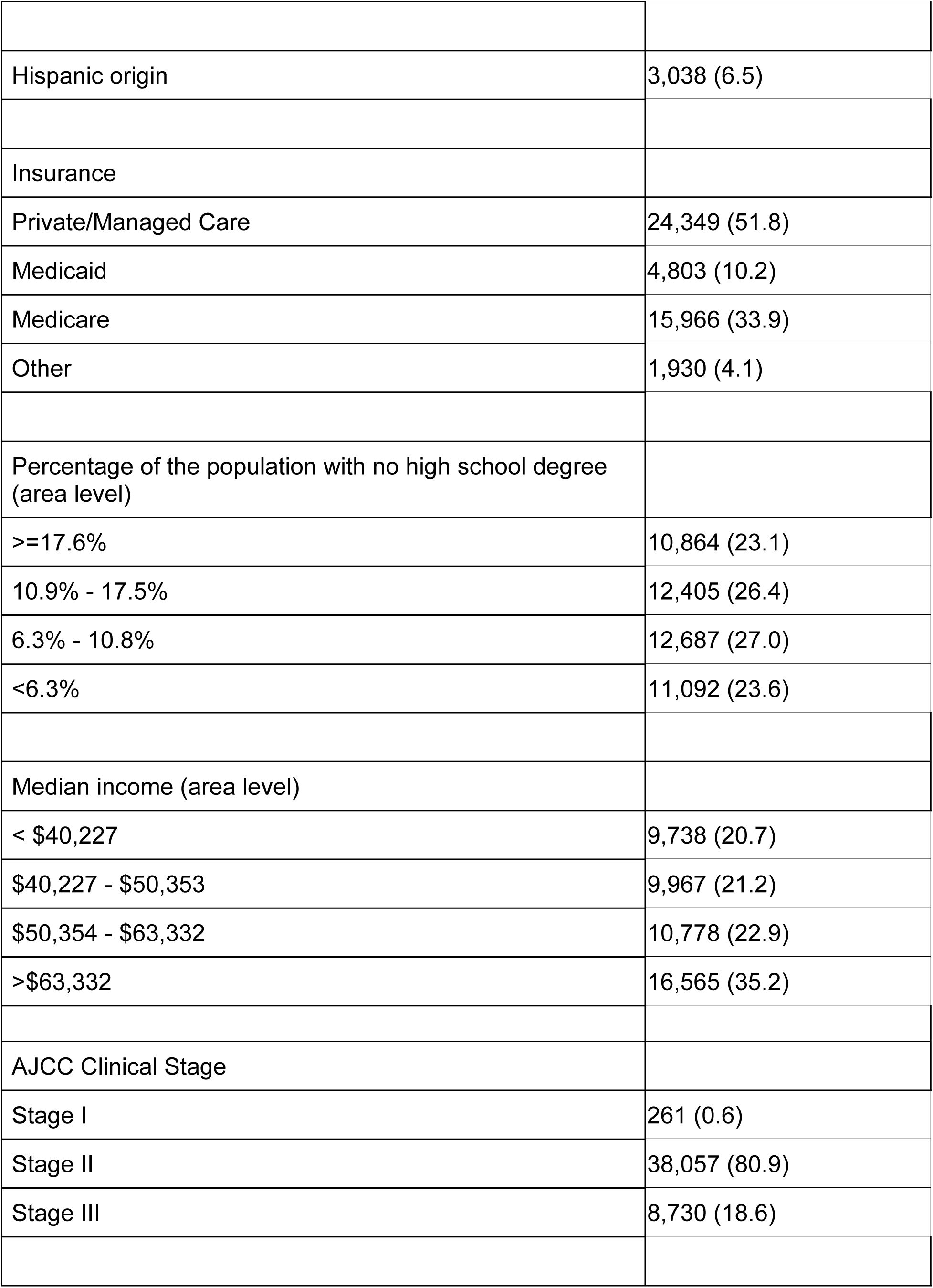
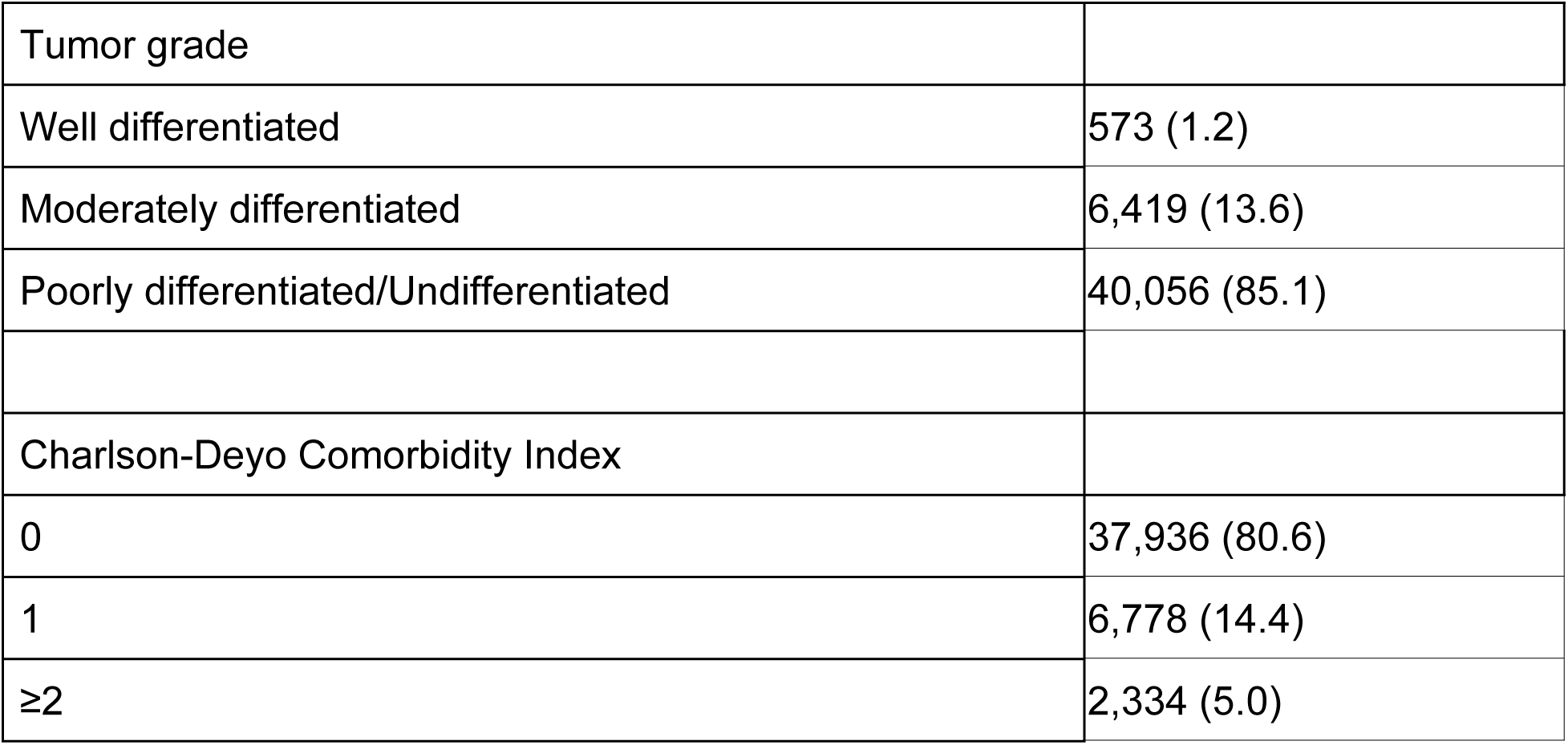
Sociodemographic and clinical characteristics of TNBC patients in NCDB (n=47,048)

Of the patients with TNBC in the OSU registry, a total of 281 patients met inclusion criteria and did not have missing data. Most patients with TNBC (Table 1) were between 45 – 64 years old (52.0%), identified as White (75.4%), and had private insurance or managed care (51.8%). Nearly half the patients lived in neighborhoods where more than 10.9% of the population had less than high-school degrees (49.5%) and over 60% had median incomes were <$50,353 (63.7%). Most patients with TNBC had low CCI scores of 0 (67.6%), had stage II disease (80.9%), and moderately differentiated tumors (68.3%). Overall, 11.0% had rapid relapse.

The characteristics of the clinical validation sample from the OSU cancer registry are described in Table 2.

| Variable | N (%) |
| --- | --- |
| Rapid Relapse | 31 (11.0) |
| Age group |  |
| <45 years | 66 (23.5) |
| 45-54 years | 75 (26.7) |
| 55-64 years | 71 (25.3) |
| 65-74 years | 48 (17.1) |
| 75+ years | 21 (7.5) |
| Race |  |
| Black | 62 (22.1) |
| White | 212 (75.4) |
| Other | 7 (2.5) |
| Hispanic origin | 4 (1.4) |
| Insurance |  |
| Private/Managed Care | 159 (56.6) |
| Medicaid | 50 (17.8) |
| Medicare | 68 (24.2) |
| Other | 4 (1.4) |
| Percentage of the population with no high school degree<br>(area level) |  |
| >=17.6% | 10,864 (23.1) |
| 10.9% - 17.5% | 12,405 (26.4) |
| 6.3% - 10.8% | 12,687 (27.0) |
| <6.3% | 11,092 (23.6) |
| Median income (area level) |  |
| < \$40,227 | 113 (40.2) |
| \$40,227 - \$50,353 | 66 (23.5) |
| \$50,354 - \$63,332 | 42 (15.0) |
| >\$63,332 | 60 (21.4) |
| AJCC Clinical Stage |  |
| Stage I | 261 (0.6) |
| Stage II | 38,057 (80.9) |
| Stage III | 8,730 (18.6) |
| Tumor grade |  |
| Well differentiated | 37 (13.2) |
| Moderately differentiated | 192 (68.3) |
| Poorly differentiated/Undifferentiated | 52 (18.5) |
| Charlson-Deyo Comorbidity Index |  |
| 0 | 190 (67.6) |
| 1 | 47 (16.7) |
| ≥2 | 44 (15.7) |

Table 3 summarizes the performance of the base models, models with SMOTE applied, and models with hyperparameter tuning after training and validating them using the NCDB data.

**Table 3:**
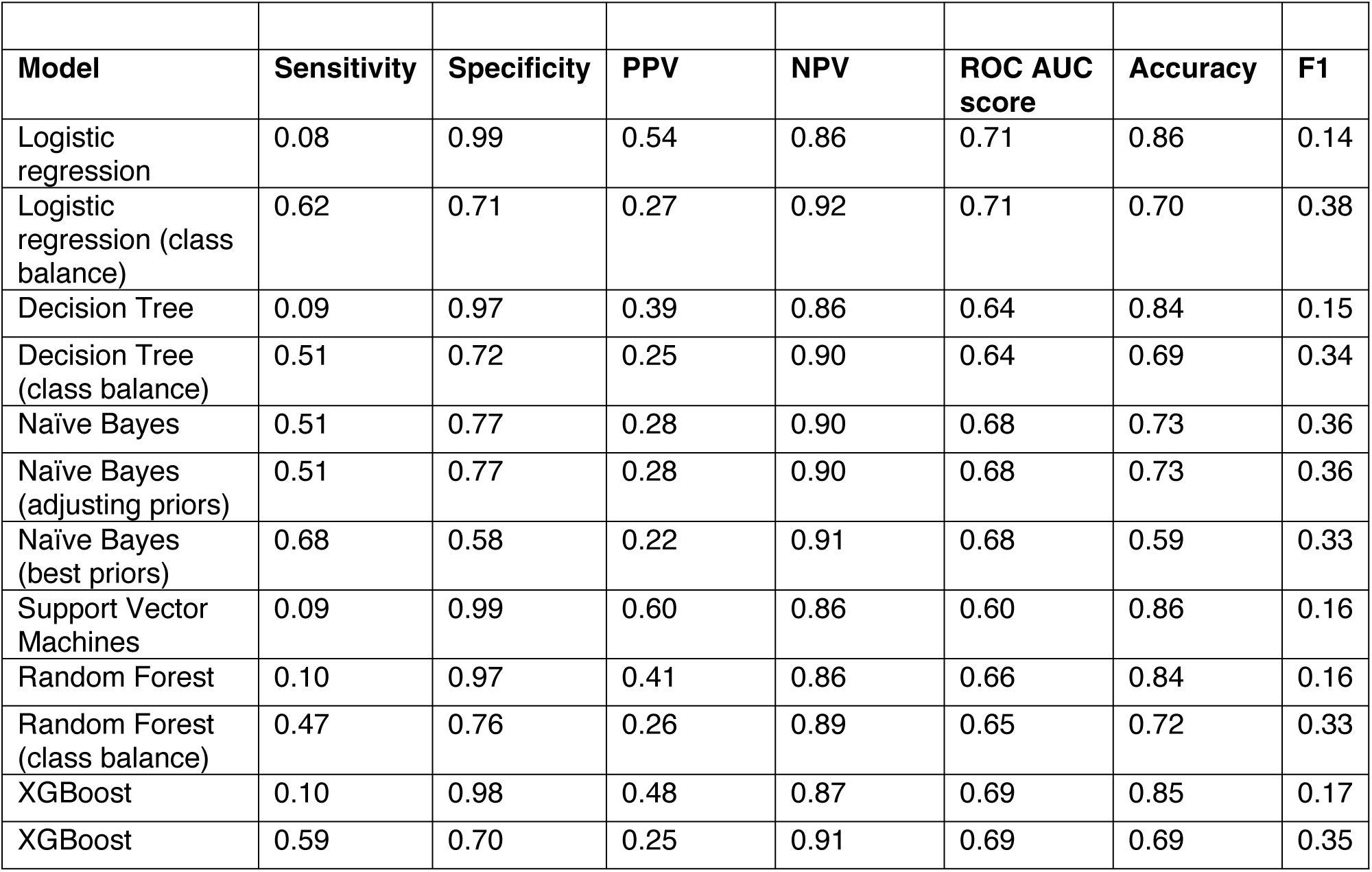
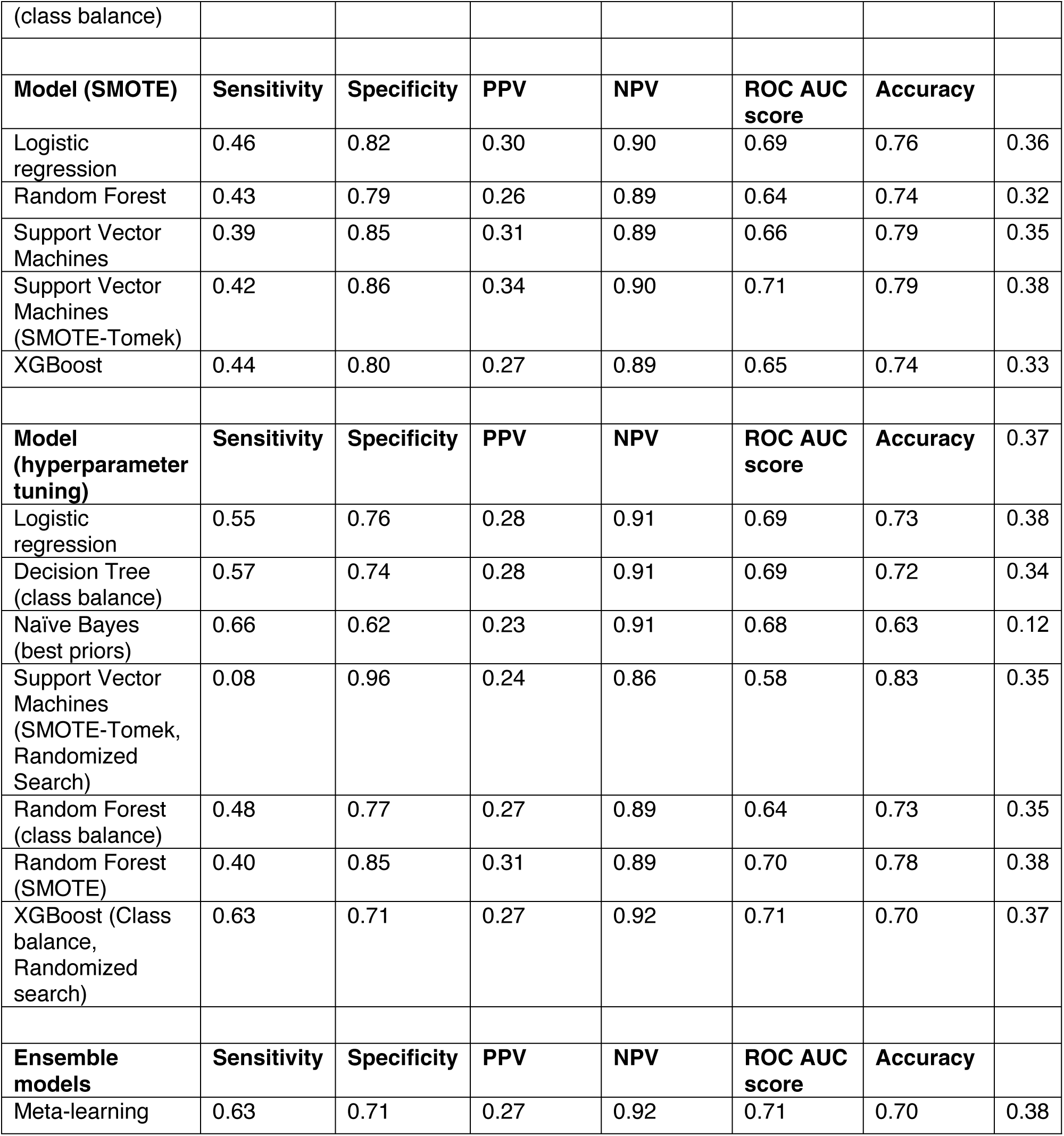
Model Performance using baseline models, models with SMOTE applied, models with hyperparameter tuning, and ensemble models trained and validated using NCDB data.

### Models trained and tested using the NCDB data

#### Baseline Models

Without data balancing or tuning, logistic regression exhibited high specificity (0.99) and low sensitivity (0.08), resulting in an accuracy of 0.86. Decision tree and random forest models exhibited moderate performance with accuracies of 0.84 each. XGBoost and SVM models demonstrated lower sensitivity of 0.10 and 0.09 respectively. Balancing the classes in logistic regression improved sensitivity (0.62) but decreased specificity (0.71), with an accuracy of 0.70. Similarly, balancing the classes in XGBoost increased the sensitivity to 0.59, decreased the specificity to 0.70, with an accuracy of 0.69.

Among the baseline models, naïve Bayes (best priors) achieved the highest sensitivity (0.68) among baseline models, but PPV (0.22) and F1 (0.33) remained low.

#### SMOTE Application

Applying SMOTE generally increased sensitivity across models, with some trade-offs in specificity. Logistic regression with SMOTE achieved a sensitivity of 0.46, specificity of 0.82, and F1 of 0.36. Random forest and SVM models also saw improved sensitivities (0.39–0.43), with F1 scores ranging from 0.32 to 0.35. The SVM-SMOTE-Tomek variant slightly improved F1 to 0.38. XGBoost saw similar modest gains.

#### Hyperparameter Tuning

Performing hyperparameter tuning influenced performance trends observed with SMOTE, as well as with baseline models. Logistic regression maintained an accuracy of 0.73 and F1 0.38. Decision tree, random forest, and naïve Bayes models showed increased sensitivity ranging from 0.48 to 0.66, and improved F1 scores to 0.35 and 0.38, respectively . However, hyperparameter tuning decreased the sensitivity to 0.08 for SVMs (F1, 0.12).

#### Overall Performance

Across all models and treatments, the ROC AUC scores ranged from 0.59 to 0.86, indicating limited discriminative ability. The models generally exhibited limited ability in identifying rapid relapse as indicated by low sensitivity values. The high NPV across all models indicates a strong ability to correctly identify non rapid relapse, while the low PPV values across many models denote a poor ability to correctly identify rrTNBC. F1 scores were generally low across models (0.12–0.38), with the highest values observed for class-balanced and ensemble approaches, indicating modest gains in overall relapse classification performance compared to baseline models.

#### Ensemble model

A meta-learning ensemble was trained using logistic regression with classes balanced, decision tree, naïve bayes with best priors, random forest with classes balanced, and XGBoost with classes balanced. The meta-learning ensemble method achieved a sensitivity of 0.63, specificity of 0.71, an AUC of 0.71, and F1 0.38.

### Testing using clinical data from the OSU registry

Table 4 presents the performance of multiple machine learning models on clinical data.

**Table 4.** Performance of models trained on NCDB and tested on real-world clinical data from the OSU registry.

| **Model** | **Sensitivity** | **Specificity** | **PPV** | **NPV** | **ROC AUC score** | **Accuracy** | **F1** |
| --- | --- | --- | --- | --- | --- | --- | --- |
| Logistic regression | 0.13 | 0.99 | 0.80 | 0.90 | 0.71 | 0.90 | 0.22 |
| Logistic regression (class balance) | 0.55 | 0.72 | 0.20 | 0.93 | 0.71 | 0.70 | 0.29 |
| Decision Tree | 0.29 | 0.95 | 0.41 | 0.92 | 0.62 | 0.88 | 0.34 |
| Naïve Bayes | 0.45 | 0.76 | 0.19 | 0.92 | 0.68 | 0.72 | 0.27 |
| Support Vector Machines | 0.10 | 0.99 | 0.60 | 0.90 | 0.62 | 0.89 | 0.17 |
| Random Forest | 0.19 | 0.96 | 0.40 | 0.91 | 0.62 | 0.88 | 0.26 |
| XGBoost | 0.13 | 0.99 | 0.67 | 0.90 | 0.66 | 0.90 | 0.22 |
| XGBoost (class balance) | 0.48 | 0.73 | 0.18 | 0.92 | 0.64 | 0.70 | 0.26 |
| **Model (SMOTE)** | **Sensitivity** | **Specificity** | **PPV** | **NPV** | **ROC AUC score** | **Accuracy** |  |
| Logistic regression | 0.42 | 0.86 | 0.27 | 0.92 | 0.70 | 0.81 | 0.33 |
| Logistic regression (class balance) | 0.42 | 0.86 | 0.27 | 0.92 | 0.70 | 0.81 | 0.33 |
| Decision Tree (class balance) | 0.45 | 0.72 | 0.17 | 0.91 | 0.61 | 0.69 | 0.25 |
| Naïve Bayes (adjusting priors) | 0.45 | 0.76 | 0.19 | 0.92 | 0.68 | 0.72 | 0.27 |
| Naïve Bayes (best priors) | 0.61 | 0.62 | 0.17 | 0.93 | 0.68 | 0.62 | 0.27 |
| Support Vector Machines | 1.00 | 0.00 | 0.11 | nan | 0.66 | 0.11 | 0.20 |
| Support Vector Machines (SMOTE-Tomek) | 0.32 | 0.87 | 0.24 | 0.91 | 0.66 | 0.81 | 0.27 |
| Random Forest (class balance) | 0.32 | 0.80 | 0.16 | 0.90 | 0.57 | 0.74 | 0.21 |
| Random forest | 0.35 | 0.80 | 0.18 | 0.91 | 0.56 | 0.75 | 0.24 |
| XGBoost | 1.00 | 0.00 | 0.11 | nan | 0.50 | 0.11 | 0.20 |
| **Model (hyperparameter tuning)** | **Sensitivity** | **Specificity** | **PPV** | **NPV** | **ROC AUC score** | **Accuracy** |  |
| Logistic regression | 0.42 | 0.86 | 0.27 | 0.92 | 0.70 | 0.81 | 0.33 |
| Decision Tree (class balance) | 0.52 | 0.80 | 0.24 | 0.93 | 0.72 | 0.77 | 0.33 |
| Naïve Byes (with best priors) | 0.58 | 0.67 | 0.18 | 0.93 | 0.68 | 0.66 | 0.27 |
| Support Vector Machines (SMOTE-Tomek) | 0.32 | 0.77 | 0.15 | 0.90 | 0.55 | 0.72 | 0.20 |
| Random Forest (class balance) | 0.39 | 0.80 | 0.20 | 0.91 | 0.61 | 0.76 | 0.26 |
| Random Forest (SMOTE) | 0.35 | 0.85 | 0.23 | 0.91 | 0.69 | 0.80 | 0.28 |
| XGBoost (class balance, randomized search) | 0.61 | 0.73 | 0.22 | 0.94 | 0.70 | 0.72 | 0.32 |
| **Ensemble models** | **Sensitivity** | **Specificity** | **PPV** | **NPV** | **ROC AUC score** | **Accuracy** |  |
| Meta-learning | 0.58 | 0.70 | 0.20 | 0.93 | 0.71 | 0.69 | 0.30 |

#### Baseline Models

The logistic regression model achieved a high specificity (0.99) with a low sensitivity (0.13) resulting in an F1 score of 0.22. Class balancing improved sensitivity to 0.55, with specificity decreased to 0.72 and an F1 of 0.29. Decision trees and random forests showed similar specificity (0.95 and 0.96, respectively) but moderate sensitivity (0.29 and 0.19, respectively) with F1 scores of 0.34 and 0.26. XGBoost models exhibited similar trends, class balancing improved sensitivity (0.13 to 0.48) and decreased specificity yielding an F1 of 0.26.

#### SMOTE

The application of SMOTE resulted in increased sensitivity across most models. Logistic regression with SMOTE showed a sensitivity of 0.42, specificity of 0.86, and F1 of 0.33. Decision trees with class balancing had sensitivity of 0.45 and specificity of 0.72 (F1 0.25). Notably, both SVM and XGBoost with standard SMOTE achieved a sensitivity of 1.00 and specificity 0.00.

#### Hyperparameter Tuning

Hyperparameter tuning influenced the performance of some models. The decision tree with class balancing exhibited a sensitivity of 0.52, specificity of 0.80, and F1 of 0.33 . XGBoost (class balance with randomized search) achieved a sensitivity of 0.61, a specificity of 0.73, and F1 of 0.32.

#### Ensemble Model

The ensemble approach using meta-learning, achieved a sensitivity of 0.58 and specificity of 0.70, with an overall ROC AUC of 0.71, accuracy of 0.69, and an F1 score of 0.30.

#### Overall Performance

Across all models and treatments, the ROC AUC scores ranged from 0.55 to 0.72, indicating limited discriminative ability. The models generally showed limited ability in identifying rrTNBC (low sensitivity). High NPV values suggest strong ability to correctly identify non-rapid relapse, while low PPV and moderate F1 scores highlight difficulty in achieving both high sensitivity and precision for the prediction of rrTNBC.

#### Finetuning models using OSU Registry data

Table 5 presents the performance of the ensemble model on the OSU registry data after applying transfer learning and threshold optimization. The fine-tuned ensemble model identified rapid relapse perfectly, with an F1 score of 0.68, although the PPV remained moderate at 0.52. After threshold optimization, the model achieved a substantially improved F1 score of 0.88, reflecting a strong balance between sensitivity (0.87) and PPV (0.90). The optimized model also demonstrated high accuracy (0.98), indicating correct predictions for most cases, and a high ROC AUC (0.99), indicating excellent discrimination between rapid relapse and non–rapid relapse.

**Table 5:** Performance of the fine-tuned, and optimized ensemble model on real-world clinical data.

| Model | Sensitivity | Specificity | PPV | NPV | ROC AUC score | Accuracy | F1 |
| --- | --- | --- | --- | --- | --- | --- | --- |
| Ensemble model (transfer learning) | 1.00 | 0.88 | 0.52 | 1.00 | 0.99 | 0.90 | 0.68 |
| Ensemble model (transfer learning + threshold optimization) | 0.87 | 0.99 | 0.90 | 0.98 | 0.99 | 0.98 | 0.88 |

## Discussion

In this study, we developed algorithms to predict rapid relapse TNBC, by training the models on the NCDB dataset and subsequently fine-tuning and testing using EHR data from the OSU cancer registry. We found the ML models trained on NCDB data tended to have high NPV or that the algorithm had excellent performance in predicting those who were not at risk of rapid relapse. Upon further finetuning the ensemble model to real-world clinical data yielded a model with high sensitivity and PPV.

Previous work suggests that receiving both surgery and chemotherapy reduces patients’ risk of rrTNBC by four times.(18, 19) While the combination of surgery and chemotherapy is guideline-concordant, research shows that there are gaps in receiving both treatments. Therefore, it is important to identify patients at risk, so every effort can be made to connect them with available resources to successfully complete treatment. Predictive tools such as CTS-5, which estimates distant recurrence risk in hormone receptor-positive breast cancer, illustrate the potential for clinical risk calculators to guide therapy decisions.(20) The rrPM leverages real-world EHR data and is tailored for TNBC, highlighting the potential for disease-specific, data-driven risk prediction. The rrPM with a sensitivity of 0.87 and a PPV of 0.90 can identify individuals at risk for rrTNBC correctly, and when it predicts relapse, it does so correctly 90% of the time.

Given the high-risk patients identified by the rrPM, linking predictions with targeted clinical navigation may help ensure timely treatment and follow-up. Patient navigation has a well-established evidence base; navigators can help overcome systemic, socioeconomic, and patient-level barriers to care, helping patients access diagnostic services, start treatment more promptly, and complete recommended care.(21, 22) By pairing rrPM risk stratification with navigation services, it may be possible to improve adherence to guideline-concordant therapy, reduce delays, and ultimately improve outcomes in rapid-relapse TNBC.

Our study adds to the limited knowledge base on predicting rapid relapse in TNBC. By focusing on a parsimonious set of data elements that are typically collected as part of routine clinical care, the rrPM can be applied to a broader patient population since fewer data requirements mean more patients meet the necessary criteria. This also facilitates adoption in resource-constrained settings as a standalone tool. Several of our models are comparable to and even outperform similar models in extant literature which use additional biomarker data requiring more testing. (5, 14)

Across the models, a key finding was the consistent trade-off between sensitivity and specificity. While some models achieved high specificity, indicating a strong ability to identify true negatives, they often had very low sensitivity, failing to detect true positives. The high NPV observed in the initial models trained on NCDB data may be particularly valuable in a clinical setting, where ruling out relapse is crucial to reduce unnecessary interventions and alleviate patient anxiety. However, the low sensitivity means that a significant portion of patients who will experience rapid relapse may be missed, which has serious implications for timely intervention. This highlights the challenge of accurately identifying those at risk for rapid relapse within our dataset.

Addressing class imbalance using approaches like SMOTE improved model performance particularly in terms of sensitivity. However, this improvement was accompanied by a decrease in specificity. While SMOTE addresses class imbalance, the introduction of synthetic data points can potentially distort the underlying data distribution. This distortion may lead to overfitting or reduced generalizability, especially in datasets with complex relationships between features. Hyperparameter tuning yielded minimal improvements in overall model performance, suggesting that limitations in predictive accuracy may stem from inherent characteristics of the data or the chosen model architectures, rather than suboptimal parameter settings. The consistently low PPV across all models has implications for its clinical applications. Overall, the models were much better at ruling out rrTNBC than they were at correctly identifying it. Low to moderate ROC AUC scores (ranging from 0.59 to 0.86) further emphasize the limited discriminative ability of the models. This suggests that the features used may not be sufficiently informative to reliably distinguish between rapid and non-rapid relapse.

The performance of the ensemble model remained suboptimal for clinical application, suggesting that the model, while showing some generalizability, may not have fully captured the nuances of the clinical data. Therefore, we employed transfer learning, cross-validation, and threshold optimization to adapt it to the registry clinical data. Indeed, this approach vastly improved the performance of the model, which is highly accurate with near perfect ROC AUC. The substantial improvement achieved through transfer learning highlights the importance of leveraging domain-specific knowledge. By fine-tuning a model pre-trained on a related dataset, we effectively transferred learned representations that were relevant to the clinical context, leading to improved performance. Further, threshold optimization allowed us to tailor the model’s decision boundary to prioritize specific clinical objectives. In this case, optimizing for high sensitivity and PPV was crucial to ensure that the model effectively identified patients at risk of relapse while minimizing false positives. This approach underscores the potential of transfer learning in adapting models to real-world clinical data, which often exhibits unique characteristics.

Our fine-tuned and optimized model for the real-world setting outperforms extant models. This study leverages training of structured data from NCDB followed by fine-tuning on real-world clinical data. In this manner, our study assessed the performance of multiple ML models and combined the best performing ones into an ensemble model in predicting rapid relapse. We then applied transfer learning to account for the nuances in real-world clinical data. Moreover, we used cross-validation during the fine-tuning process to maximize efficiency of the available data as well as to mitigate any overfitting owing to the small sample size of the clinical data, thereby improving its robustness. As such the rrPM may complement existing screening and diagnostic tests providing critical insights in treatment outcomes to potentially improve outcomes in TNBC care.

Additionally, compared to other standard tests which may subject patients to some risks (e.g. radiation), our model presents no bodily risk to the patient. Another key strength is testing on clinical data to assess how well the model generalizes to completely new data. Moreover, the rrPM prioritized features that could eventually be easily mapped to Fast Healthcare Interoperability Resources (FHIR). This was done to facilitate easier translation since these features could be easily extracted using application programming interfaces (APIs) requiring minimal staging or processing. By integrating rrPM into clinical workflows, automated alerts or dashboards could facilitate navigation of at-risk patients, supporting oncologists, nurses, and patient navigators in targeting interventions efficiently. This clinical navigation potential extends the model’s utility beyond prediction, toward actionable decision support in TNBC care.

Despite the promising results, our study has limitations. The relatively small sample size of the clinical dataset may limit the generalizability of the model. Therefore, further validation of the model on larger, independent datasets and exploring external validation are warranted. Additionally, investigating the model’s performance across different patient subgroups could provide valuable insights into its clinical utility.

Additionally, clinical validation is crucial to determine the true utility of rrPM in predicting relapse. Prospective studies that evaluate the model’s impact on clinical decision-making and patient outcomes are essential to ensure its safe and effective implementation. Future implementation of rrPM could be coupled with clinical navigation programs to prioritize high-risk patients for support services, monitoring, and follow-up, ensuring that predictive insights translate into improved patient outcomes.

In summary, we developed and optimized machine learning models to predict cancer relapse using structured and real-world clinical data. Initial models exhibited high negative predictive value but low sensitivity. Transfer learning, cross-validation, and threshold optimization significantly improved performance, yielding a model with high sensitivity and positive predictive value. This optimized model, utilizing readily available clinical data and potentially translatable to FHIR, demonstrates promise for identifying high-risk patients, though clinical validation is essential.

## Materials and Methods

### Data sources

#### National Cancer Database (NCDB)

The NCDB is a joint program of the American College of Surgeons, Commission on Cancer (CoC), and the American Cancer Society. The models are based on the 2019 Participant User File comprising patients diagnosed with TNBC between 2010 and 2019. Patients of TNBC were identified based on the absence of ER, PR, and HER2. To train and validate our ML model, we identified patients diagnosed with TNBC between 2010 and 2019 with American Joint Commission on Cancer (AJCC) clinical stage IB to IIIC, who had at least 24 months of follow-up in the NCDB. We excluded stage IA and IV, as well as inflammatory cancers from our analysis since they are inherently different in terms of treatment guidelines and prognosis. Patients who were alive, but did not have at least 24 months of follow-up for any reason were excluded.

#### The Ohio State University Cancer Registry

For testing the algorithm, we identified all patients with TNBC diagnosed between 2012 and 2020, receiving care at the James Comprehensive Cancer Center identified through the OSU Cancer Registry. Briefly, the registry had information on 4,492 patients with breast cancer. Of these, 640 patients were diagnosed with TNBC based on the absence of ER, PR, and HER2. We included patients 18 and over, with TNBC, and AJCC clinical stage IB to IIIC. We also excluded those with inflammatory cancers since they are different in terms of treatment guidelines and prognosis. Thus, our final testing dataset had 281 patients.

#### Outcome Definition

The prediction target was rapid relapse or rrTNBC, defined as all-cause mortality within 24 months of diagnosis. This operational definition is consistent with prior large-scale TNBC studies.(3, 23–26)

#### Input Features

The model used a structured set of features, drawn from patient-level and community-level data:

- Sociodemographic features: age, race, Hispanic ethnicity, insurance type, and marital status.
- Community needs: census tract-level metrics from the American Community Survey, including percent of adults without a high school diploma and median household income.
- Clinical features: AJCC clinical stage, tumor grade (differentiation), and the Charlson Comorbidity Index (CCI), used to capture comorbidity burden.

#### Model Development

We framed the prediction of rapid relapse in TNBC as a supervised binary classification task (rapid relapse vs. no rapid relapse).

#### Data Partitioning Strategy

The full dataset from NCDB was partitioned into training (80%) and testing (20%) sets using stratified random sampling to preserve the proportion of rapid relapse vs. non-relapse. The training subset was used for all model development, including preprocessing, imbalance handling, hyperparameter tuning, and model selection. The held-out 20% test set was not accessed during those steps and served only for final evaluation. Within the training set, a 5-fold stratified cross-validation was applied for hyperparameter tuning and model comparison. Each fold maintained the class balance observed in the training data. All operations that could leak information —feature scaling and resampling— were fit only on the training folds and then applied to the validation fold, to avoid optimistic bias.

#### Feature Engineering and Preprocessing

All candidate predictors available at diagnosis were included: sociodemographic factors, clinical staging, comorbidity scores, tumor grade, insurance, area-level SDoH. No *a priori* feature elimination was done; instead, model performance and regularization were expected to down-weight irrelevant variables. Categorical features (race, insurance, marital status, area-level categories) were encoded using ordinal encoding so that all models (tree-based and linear) could accept them without one-hot explosion.

Tree-based models (decision tree, random forest, XGBoost) worked on unscaled features, because they are invariant to monotonic transformations.

#### Handling Class Imbalance

Rapid relapse was a minority class (∼14-15% in NCDB). We addressed this through:

1. Baseline (no class adjustment) to assess raw model performance.
2. Algorithmic class weighting, where supported (e.g., logistic regression, decision tree, random forest), using class_weight=’balanced’ to penalize misclassification of relapse more heavily.
3. Data-level resampling with SMOTE + Tomek Links via SMOTETomek() to oversample the rapid relapse class and remove overlapping majority samples that lie near class boundaries. This hybrid approach was adopted in models where class weighting was insufficient or inappropriate.

Each model type was trained under all three conditions (no balancing, class weighting, SMOTE-Tomek), so that the effect of imbalance correction could be compared per algorithm.

#### Model Types, Hyperparameter Tuning, and Evaluation

Six machine learning models were trained and evaluated: 1. Logistic Regression; 2. Decision Tree Classifier; 3. Random Forest Classifier; 4. Gaussian Naïve Bayes; 5. Support Vector Machine (SVM); and 6. XGBoostClassifier Hyperparameter optimization was conducted using RandomizedSearchCV with 5-fold stratified cross-validation nested within the training set. Parameter grids included:

- Logistic regression: regularization parameter C over ranges (e.g., 1e-4 to 1e4), penalty type as L2.
- Decision tree: max_depth, min_samples_split, min_samples_leaf.
- Random forest: n_estimators, max_depth, min_samples_leaf, max_features.
- Support Vector Machines (SVM): kernel type (linear, RBF), C, and where appropriate, gamma.
- XGBoost: learning_rate, n_estimators, max_depth, subsample, colsample_bytree.

The primary selection criterion during tuning was ROC AUC, but models were also compared using F1 score, sensitivity, specificity, precision or positive predictive value (PPV), negative predictive value (NPV), and accuracy. After tuning, each model was retrained on the full training set using its best hyperparameters.

#### Ensemble Modeling

Two ensemble strategies were constructed to capitalize on model diversity:

- A soft voting ensemble, combining probabilistic outputs from logistic regression, decision tree, and random forest. The average probability across these base learners was used for final class prediction, which helped produce smoother calibrated predictions and moderate overconfident outputs from individual models.
- A stacked ensemble, using logistic regression as a meta-learner. Base learners were the same three (logistic regression, decision tree, random forest). During cross-validation, out-of-fold predictions from base models were generated to form meta-features for the second layer. This approach was intended to let the meta-learner learn patterns in how and when different base learners erred, thereby improving overall prediction.

The stacking model was chosen as the final ensemble because it achieved the best tradeoff among sensitivity, F1 score, and ROC AUC in internal validation.

#### Threshold Setting and Transfer Learning to OSU Data

To improve performance in a local clinical setting, the final stacking ensemble model—originally trained on the NCDB data—was adapted to institutional data from OSU Cancer Registry. This step constitutes a form of transfer learning, wherein knowledge from a large, diverse national dataset was transferred to a smaller, institution-specific dataset with the same structure but potentially different distributions and prevalence patterns.

The pretrained NCDB model was applied to the OSU dataset using 5-fold stratified cross-validation. This allowed evaluation of how well the model generalized to local data and highlighted potential domain shifts. Based on the performance metrics observed across these folds—particularly calibration and precision-recall tradeoffs—the decision threshold for classifying a patient as high-risk was adjusted. The optimal probability cutoff was selected to maximize the F1 score, balancing sensitivity and precision, while ensuring positive predictive value remained clinically acceptable.

This transfer learning approach allowed the model to retain the generalizability conferred by the NCDB data, while tuning its decision boundary to better fit local practice patterns, patient demographics, and base relapse rates.

To improve real-world performance, the stacking model trained on NCDB data was fine-tuned using the OSU Cancer Registry data. First, the model was applied to the OSU data in cross-validation mode (again with 5 folds stratified), to measure calibration and detection performance under local distributions. Then the classification threshold was adjusted (not just default 0.5) by exploring probability prediction thresholds that maximize F1 score while also keeping precision/PPV high enough for clinical utility. This threshold tuning was done using ROC and precision-recall curve analyses on the OSU validation folds, to reflect local prevalence and acceptable tradeoffs.

#### Model evaluation

All models were evaluated on the held-out test set using the following metrics:

- Sensitivity (Recall): proportion of rrTNBC cases correctly identified
- Specificity: proportion of non-rrTNBC cases correctly identified
- Precision (Positive Predictive Value): proportion of predicted rrTNBC cases that were correct
- Negative Predictive Value: proportion of predicted non-rrTNBC cases that were correct
- Accuracy: proportion of all cases (both rrTNBC and non-rrTNBC) that were correctly identified
- Area under the ROC curve (ROC AUC): a measure of a model’s ability to distinguish between rrTNBC and non-rrTNBC cases at various threshold settings; it represents the overall performance of a classification model. Where appropriate, we also examined the precision-recall curve, given the class imbalance.
- F-1 score: the harmonic mean of the model’s ability to correctly identify patients with rrTNBC (sensitivity) and the proportion of predicted rrTNBC that are truly rapid relapse (precision or PPV).

Performance of all final models (baseline, class-weighted, SMOTE/Tomek, hyperparameter-tuned, and ensemble) was reported on the held-out test set from NCDB and on the OSU dataset after fine-tuning. The held-out test set was never used during any training, tuning, or threshold optimization, ensuring no data leakage. Model performance was assessed both before and after threshold tuning on institutional data, enabling direct comparison between general model behavior and deployment-calibrated behavior. The OSU dataset was used for testing even when initial model performance was suboptimal to evaluate generalizability and real-world applicability.

#### Transfer Learning and Model Fine-Tuning

To further improve model performance and adapt it to real-world clinical data, we applied transfer learning by fine-tuning the final ensemble model on the OSU dataset. This involved re-training the model using stratified k-fold cross-validation to ensure balanced evaluation across folds. We also optimized the classification threshold to improve the balance between recall (sensitivity) and precision (PPV), based on predicted probabilities. This approach allowed the model to better generalize to the clinical dataset and improve relevance in a real-world setting.

#### Software and Tools

Data preprocessing and descriptive analyses were conducted in Stata/SE 17.0. All model development and evaluation were carried out using Python version 3.12.1. The analysis leveraged the scikit-learn library (v1.4.2) for model construction, xgboost (v2.0.3) for gradient boosting models, and imbalanced-learn (v0.12.2) for resampling and imbalance correction.

## Data Availability

The data underlying the results presented in the study are available from - https://www.facs.org/quality-programs/cancer-programs/national-cancer-database/puf/ and https://u.osu.edu/secondarydatacore/osu-pcornet-common-data-model-cdm/\

https://u.osu.edu/secondarydatacore/osu-pcornet-common-data-model-cdm/

